# Quantifying the Cost of Measles Outbreak in the U.S. and How Costs Scale with Outbreak Size

**DOI:** 10.1101/2025.10.24.25338724

**Authors:** Salin Sriudomporn, Bryan Patenaude

## Abstract

**Importance:** In 2025, the United States experienced the largest number of measles cases since its elimination in 2000. These outbreaks require extensive public health responses to mitigate, consume substantial hospital resources, and impose an economic and financial burden on state and federal budgets.

**Objective:** To review evidence on the costs of measles outbreaks in the United States between 2000 and 2025, as well as to examine how costs scale with outbreak size to better model the costs associated with measles outbreaks.

**Evidence Review:** We conducted a literature review using PubMed to identify studies published between January 1, 2000, and October 7, 2025, using terms related to measles cases, costs, and outbreak response, across all 50 states in the United States of America. Additional publicly available reports beyond the PubMed published literature were identified through a review of official CDC and state health department reports.

**Findings:** A total of 120 articles were screened, 33 underwent full text review, and data were extracted from 19 articles, yielding outbreak relevant costs in 18 different states. Eight additional reports in the gray literature were identified. Outbreak size ranged from 1 to 802 cases. Average outbreak cost per case was estimated at $43,203.65, while the average cost per contact was $443.46. Average cost per case varied from $33,415.75 from the medical provider perspective to $58,591.50 when including public health response costs. The incremental cost per case was estimated at $16,197.13 per additional measles case, after accounting for the fixed costs of initiating a public health response.

**Conclusion and Relevance:** Measles outbreaks in the U.S. continue to re-emerge and impose significant financial and public health burden. These outbreaks are largely preventable through inexpensive and highly effective measles vaccination paired with system-level preparedness for effective containment. Since fixed response costs for any outbreak can be costly, our estimates provide improved data on how costs of measles outbreaks scale with outbreak size by isolating incremental costs associated incident new cases, as well as incident contacts, for use alongside budgetary planning and modeled risk assessments.

## Introduction

Measles was declared eliminated in the United States in 2000.^1^ Nonetheless, outbreaks have emerged intermittently over the past two decades, as vaccination rates have declined in many communities across the U.S. In communities with low or declining coverage imported measles virus infections can be easily transmitted through contact with the unvaccinated, susceptible population and can quickly lead to an outbreak.^2^ Once an outbreak occurs, effective containment relies on rapid public health responses, including case investigation, contact tracing, quarantine, and vaccination campaigns. These measures incur substantial economic and financial costs, with large initial fixed costs and incremental costs that continue to scale as an outbreak expands.

A systematic review by Pike et al. (2020) ^3^ synthesized evidence on outbreak costs in the United States between 2004 and 2017. Since then, several large outbreaks have occurred, including 2018-2019 epidemic in New York City, and most recent widespread outbreak originating in Texas in 2025. As of October of 2025, the U.S. has recorded the largest number of measles cases since elimination was declared in 2000. As outbreak sizes have increased and immunization rates continue to decline, the risk of further larger-scale outbreaks is heightened. Expanding the evidence to cover all outbreaks between 2000-2025, this article aims to fill an evidence gap for federal, state, and local communities on how the costs of measles outbreaks are expected to scale with outbreak size. This data can enable better financial planning for outbreak response and improve mobilization of resources to mitigate further health and financial impacts.

## Methodology

We conducted a systematic review following the Preferred Reporting Items for Systematic Reviews and Meta-Analyses (PRISMA) statement guidelines (Figure 1), using PubMed to identify English-language studies published between January 1, 2000, and October 7, 2025 that contained search terms related to measles cases, costs, and outbreak response (Appendix 1). All titles and abstracts relevant to our study were retrieved and a full-text review was completed. Inclusion and exclusion criteria can be found in Appendix 2. Articles were limited to data on all 50 states within the United States of America. Additionally, the gray literature, defined as official publicly available government reports that were not indexed on PubMed, was examined and several relevant reports from the US Centers for Disease Control (CDC), state health departments, and news article citations were identified.

**Figure 1.**
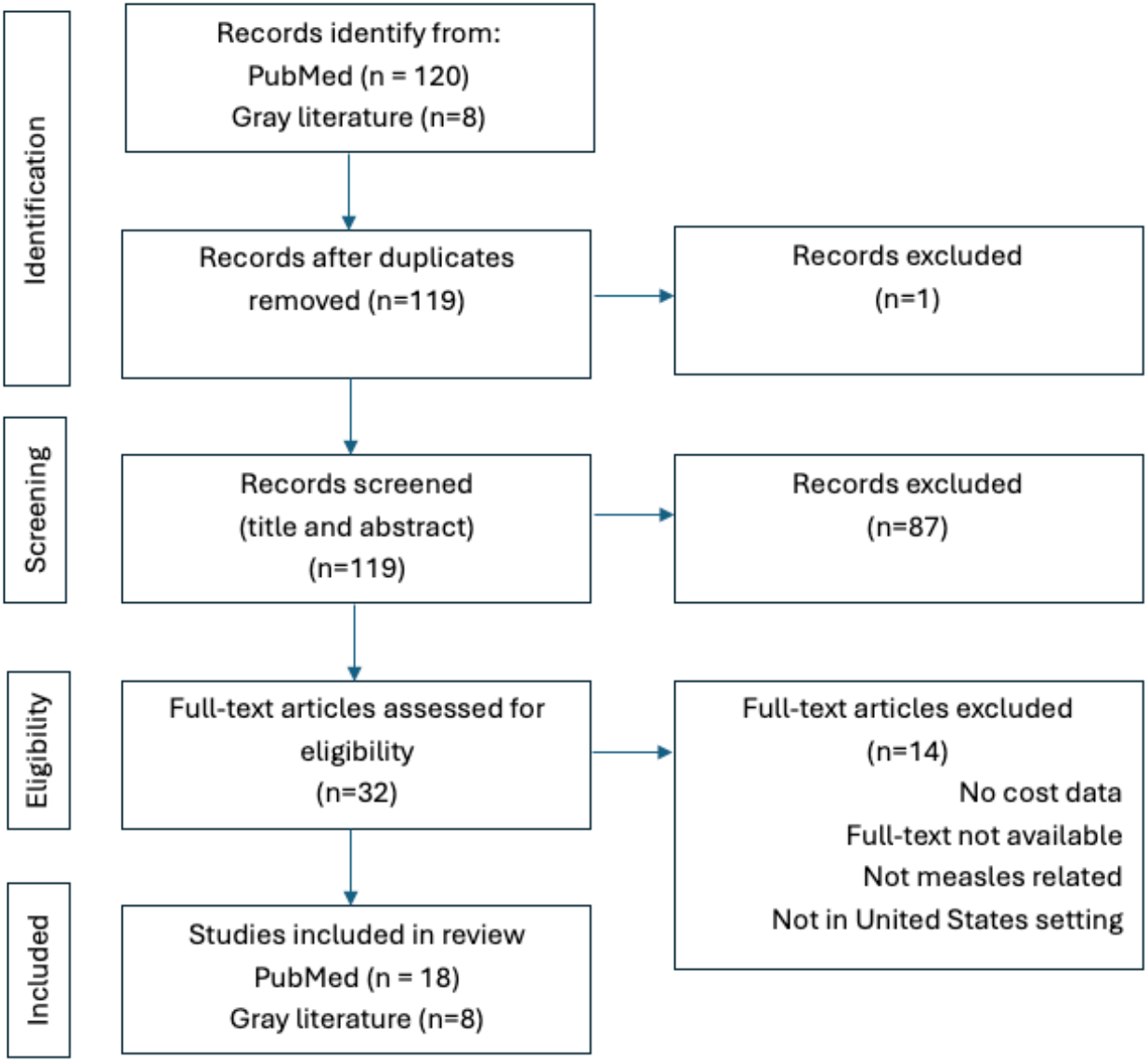
PRISMA diagram

From the data, we extracted the year of outbreak, number of cases, number of contacts, total costs, cost perspective, cost per case, and cost per contact. All costs were converted to 2024 USD in order to account for inflation and improve comparability over the defined time horizon. When possible, costs were disaggregated into fixed costs, variable costs, as well as costs incurred due to outbreak response, medical treatment, or indirect economic costs such as productivity loss.

Once data was compiled, we conducted a simple linear regression analysis to estimate the fixed and incremental cost per case to better understand how costs have historically scaled with outbreak size. Costs related to productivity loss were excluded from this calculation.

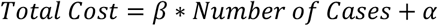

where β = Fixed cost, α = incremental cost per case

## Results

The search strategy identified 120 articles, of which 33 articles were identified as containing relevant data. A full-text review was conducted for these articles, and final metadata was extracted from 18 articles after duplicate data points were discarded (Table 1). The extracted data covered 18 states: Washington (WA), Minnesota (MN), Indiana (IN), Arizona (AZ), Illinois (IL), New York (NY), Kentucky (KY), Iowa (IA), Colorado (CO), California (CA), Michigan (MI), Texas (TX), New Hampshire (NH), North Carolina (NC), Maryland (MD), Oklahoma (OK), Massachusetts (MA), and Wisconsin (WI). Eight additional reports from the gray literature were identified. From these reports, we extracted 51 additional data points related to the number of cases, 42 of which were from the 2025 outbreak alone (Table 2).

**Table 1.** Total and Per Costs of United States Measles Outbreaks.

| **Study** | **Location** | **Year** | **Number of Cases** | **Number of Contacts** | **Total cost (2024USD)** | **Cost per Case (2024 USD)** | **Cost per Contact (2024 USD)** | **Ref.** |
| --- | --- | --- | --- | --- | --- | --- | --- | --- |
| Pike et al. (2021) | Clark County, WA | 2019 | 71 | 4011 | \$4,319,626.29 | \$59,994.20 | \$1,028.57 | 7 |
| Hester et al. (2019) | Minneapolis, MN | 2017 | 21 | | \$1,713,293.08 | \$6,973.10 | | 9 |
| Hester et al. (2019) | Minneapolis, MN | 2011 | 7 |  |  |  |  | 9 |
| Chovatiya & Silverberg (2020)* | United States | 2002-2016 | 1018 | | \$22,370,065.79 | \$9,802.67 | | 10 |
| Fiebelkorn, Seward & Orenstein (2014) | Indiana, IN | 2005 | 3 | | \$186,914.61 | | | 11 |
| Fiebelkorn, Seward & Orenstein (2014) | Arizona, AZ | 2008 | 7 | | \$1,199,051.04 | | | 11 |
| Fiebelkorn, Seward & Orenstein (2014) | Chicago, IL | 2008 | 1 | | \$28,358.21 | | | 11 |
| Rosen et al. (2018) | NYC, NY | 2013 | 58 | 3351 | \$546,992.10 | \$9,430.90 | \$163.23 | 12 |
| Zucker et al. (2020) | NYC, NY | 2018-2019 | 649 | 20000 | \$10,614,192.40 | \$16,354.69 | \$530.71 | 6 |
| Lo & Hotez (2017)* | United States | | | | \$2,767,627.28 | \$26,358.36 | | 13 |
| Wendorf et al. (2015) | King County, WA | 2013 | 1 | 122 | \$11,995.20 | \$11,995.20 | \$98.46 | 14 |
| Chen et al. (2011) | Tucson, AZ | 2008 | 14 | 8231 | \$1,368,078.64 | \$97,719.35 | \$165.88 | 15 |
| Pike, Leidner & Gastañaduy (2020) | Albany, NY | 2011 | 2 | 93 | \$98,380.29 | \$49,190.79 | \$371.62 | 3,16 |
| Marx et al. (2017) | Denver, CO | 2016 | 6 | 283 | \$63,996.24 | \$66,898.44 | \$236.60 | 16 |
| Marx et al. (2017) | Denver, CO | 2017 | 1 | 232 | \$23,689.50 | \$24,157.56 | \$104.16 | 16 |
| Ortega-Sanchez et al. (2014) | Iowa, IA and Michigan, MI | 2004 | 1 | | \$310,698.98 | | \$311.25 | 17 |
| Ortega-Sanchez et al. (2014) | California, CA | 2008 | 12 | 376 | \$195,199.00 | | \$519.15 | 17 |
| Ortega-Sanchez et al. (2014) | Kentucky, KY | 2010 | 1 | 45 | \$36,398.75 | | \$808.89 | 17 |
| Sugerman et al. (2010) | San Diego, CA | 2008 | 12 | 839 | \$302,980.78 | \$24,152.41 | \$345.90 | 18 |
| Coleman et al. (2017) | Texas, TX | 2011 | 0 | | \$3,295.98 | | | 8 |
| Coleman et al. (2017) | New Hampshire (NH) | 2011 | 0 | | \$993.82 | | | 8 |
| Coleman et al. (2017) | North Carolina, NC | 2011 | 1 | | \$12,705.69 | \$12,705.69 | | 8 |
| Coleman et al. (2017) | Maryland, MD | 2011 | 2 | | \$22,113.96 | \$11,056.98 | | 8 |
| Coleman et al. (2017) | Oklahoma, OK | 2011 | 0 | | \$891.85 | | | 8 |
| Coleman et al. (2017) | California, CA | 2011 | 4 | | \$50,430.70 | \$12,607.67 | | 8 |
| Coleman et al. (2017) | Wisconsin, WI | 2011 | 0 | | \$3,027.42 | | | 8 |
| Coleman et al. (2017) | Massachusetts | 2011 | 0 | | \$3,027.42 | | | 8 |
| Coleman et al. (2014) | Louisville, KY | 2010 | 1 | 44 | \$42,182.82 | \$42,182.82 | \$959.26 | 19 |
| Marx et al. (2017) | Arapahoe County,<br>Colorado, USA | 2016-2017 | 2 | 31 | \$65,591.45 | \$32,795.72 | \$187.14 | 16 |
| Dayan et al. (2005) | Iowa | 2004 | 1 | 1000 | \$243,614.79 | \$243,614.79 | \$242.84 | 5 |
| Parker et al. (2006) | Indiana | 2005 | 34 | 500 | \$277,369.71 | \$8,157.93 | \$554.74 | 20 |
| McCullough et al.<br>(2019) | Phoenix, AZ | 2015 | 2 | 215 | \$195,847.61 | \$97,923.80 | \$910.39 | 21 |
| Average | | | | | \$756,584.08 | \$43,203.65 | \$443.46 | |

\* Excluded from the total cost average and perspective analysis as the studies contain more than one outbreak

**Table 2.** Total and Per Costs of United States Measles Outbreaks by Perspective.

| **Study** | **Location** | **Total cost**<br>**(2024USD)** | **Cost per Case**<br>**(2024 USD)** | **Cost per Contact**<br>**(2024 USD)** |
| --- | --- | --- | --- | --- |
| ***Provider Perspective*** | | \$556,583.01 | \$33,415.75 | \$512.87 |
| Pike et al. (2021) <sup>7</sup> | Clark County,<br>WA | \$95,618.71 | \$1,328.04 | - |
| Hester et al. (2019) <sup>9</sup> | Minneapolis, MN | \$1,713,293.08 | \$6,973.10 | - |
| Fiebelkorn, Seward & Orenstein (2014) <sup>11</sup> | Indiana | \$186,914.61 | - | - |
| Fiebelkorn, Seward & Orenstein (2014) <sup>11</sup> | Arizona | \$1,199,051.04 | - | - |
| Fiebelkorn, Seward & Orenstein (2014) <sup>11</sup> | Chicago | \$28,358.21 | \$28,358.21 | - |
| Chen et al. (2011) <sup>15</sup> | Tucson, AZ | \$1,368,078.64 | \$97,719.35 | \$165.88 |
| Pike, Leidner & Gastañaduy (2020) <sup>3</sup> | Louisville, KY | \$42,182.82 | \$42,182.82 | \$959.26 |
| Pike, Leidner & Gastañaduy (2020) <sup>3</sup> | Albany, NY | \$98,380.29 | \$49,190.79 | \$371.62 |
| Parker et al. (2006) <sup>20</sup> | Indiana | \$277,369.71 | \$8,157.93 | \$554.74 |
| ***Public Health Perspective*** | | \$766,013.80 | \$58,591.50 | \$377.04 |
| Pike et al. (2021) <sup>7</sup> | Clark County,<br>WA | \$2,923,291.37 | - | - |
| Rosen et al. (2018) <sup>12</sup> | NYC, NY | \$546,992.10 | \$9,430.90 | \$163.23 |
| Zucker et al. (2020) <sup>6</sup> | NYC, NY | \$10,614,192.40 | \$16,354.69 | \$530.71 |
| Wendorf et al. (2015) <sup>14</sup> | King County,<br>WA | \$11,995.20 | \$11,995.20 | \$98.46 |
| Marx et al. (2017) <sup>16</sup> | Denver, CO | \$63,996.24 | \$66,898.44 | \$236.60 |
| Marx et al. (2017) <sup>16</sup> | Denver, CO | \$23,689.50 | \$24,157.56 | \$104.16 |
| McCullough et al. (2019) <sup>21</sup> | Phoenix, AZ | \$195,847.61 | \$97,923.80 | \$910.39 |
| Ortega-Sanchez et al. (2014) <sup>17</sup> | California | \$195,199.00 | - | \$519.15 |
| Ortega-Sanchez et al. (2014) <sup>17</sup> | Kentucky | \$36,398.75 | - | \$808.89 |
| Sugerman et al. (2010) <sup>18</sup> | San Diego, CA | \$302,980.78 | \$24,152.41 | \$345.90 |
| Coleman et al. (2017) <sup>19</sup> | Texas | \$3,295.98 | - | - |
| Coleman et al. (2017) <sup>19</sup> | New Hampshire | \$993.82 | - | - |
| Coleman et al. (2017) <sup>19</sup> | North Carolina | \$12,705.69 | \$12,705.69 | - |
| Coleman et al. (2017) <sup>19</sup> | Maryland | \$22,113.96 | \$11,056.98 | - |
| Coleman et al. (2017) <sup>19</sup> | Oklahoma | \$891.85 | - | - |
| Coleman et al. (2017) <sup>19</sup> | California | \$50,430.70 | \$12,607.67 | - |
| Coleman et al. (2017) <sup>19</sup> | Wisconsin | \$3,027.42 | - | - |
| Coleman et al. (2017) <sup>19</sup> | Massachusetts | \$3,027.42 | - | - |
| Marx et al. (2017) <sup>16</sup> | Arapahoe<br>County,<br>Colorado, USA | \$65,591.45 | \$32,795.72 | \$187.14 |
| Dayan et al. (2005) <sup>3,5</sup> | Iowa | \$243,614.79 | \$243,614.79 | \$242.84 |
| ***Societal Perspective*** | | \$1,300,716.21 | 18,065.61 | \$309.58 |
| Pike et al. (2021) <sup>7</sup> | Clark County,<br>WA | \$1,300,716.21 | 18,065.61 | \$309.58 |

As of October 7, 2025, a total of 1,563 measles cases have been reported across 41 states. Cases were reported in all states except Connecticut, Delaware, Maine, Massachusetts, Mississippi, Nevada, New Hampshire, North Carolina, and West Virginia, with Texas experiencing the largest outbreak at 802 cases, and incurring an estimated $10 million in costs.^4^

Cost per case varied widely across outbreaks, depending on the state, number of cases, number of contacts, and the costing perspective taken. The earliest outbreak identified in our systematic review within the past 25 years was a single-case event in Iowa in 2004. From the public health perspective, the total cost was estimated at $243,614.79 ^5^, representing the highest cost per case due to the small outbreak size with a high proportion of fixed costs. The most detailed costing data presented come from New York City (2018–2019) and Clark County, Washington (2019). These data covered 649 cases in New York City resulting in a total public health response cost of $10.6 million, or $16,355 per case ^6^, while the Clark County outbreak involved 71 cases incurring total societal costs estimated at $4.3 million or $59,994 per case.^7^

Across all evaluations, the average total cost of a measles outbreak was estimated at $756,584.08 (range, $891.85 - $10,614,192.40). Note that the lowest reported cost of $891.85 was for Oklahoma where no cases were identified but four refugee contacts were investigated, and the total event cost was attributed to the costs of labor and measles testing expenses.^8^ The average cost per case across all studies was estimated at $43,203.65 (range, $6,973.10 - $243,614.79), while the average cost per contact was substantially lower, estimated at $443.46 (range, $98.46 - $1,028.57) (Table 1).

There are important differences in total and unit costs between costing perspectives, so cost estimates by perspective were calculated for comparison. The perspectives taken in the literature were the public health perspective, provider perspective, and societal perspective. A public health perspective typically includes direct medical costs of treatment and quarantine protocols at facilities as well as upstream costs of outbreak response and response management. A provider perspective typically includes direct medical costs incurred by healthcare providers and outbreak response measures employed by healthcare providers such as labor, personal protective equipment, and costs associated with quarantine protocols. A societal perspective incorporates both the public health perspective and the provider perspective as well as adding the patient perspective by including additional direct costs borne by the patient, such as transportation costs as well as indirect costs such as lost labor productivity due to time off work of the patient or primary caregiver and, potentially, long-term productivity losses due to disability or premature death of the patient. When examining results by perspective, the total cost from a public health perspective was estimated at $766,013.80 (range: $891.85 - $10,614,192.40). The corresponding cost per case ranged from $9,430.90 to $243,614.79, averaging $58,591.50, while the cost per contact ranged from $98.46 to $910.39, with an average of $377.04. From the provider perspective, direct medical costs incurred by facilities, averaged $556,583.01 (range, $42,182.82 - $1.7 million), with a cost per case of $33,415.75 on average (range, $1,328.04 – $97,719.35) and cost per contact of $512.87 (range, $165.88 -$959.26). Pike et al. was the only study that employed the societal perspective and included patient productivity loss in the analysis.^7^ The productivity loss from the Clark County, WA outbreak amounted to $1,300,716.21, corresponding to $18,065.61 per case and $309.58 per contact. Given that productivity loss is not directly reflected in public budgets or expenditures, the productivity loss portion of this estimate was excluded from the regression analysis so it would be comparable with the other studies. Complete results are presented in Table 2.

Our regression analysis estimated substantial fixed costs at $244,480.40 (95% CI: $21,152.01 - $467,808.79) and each additional measles case was associated with an incremental cost of $16,197.13 (95% CI: $15,609.32 - $16,784.94). Complete results are presented in Table 3. Nonlinearities were explored to estimate marginal cost but a standard linear fit best represented the data resulting in the incremental cost being equivalent to marginal cost. For small outbreaks, over 50% of costs incurred could be attributed to fixed costs, which would have been incurred at the same level regardless of the outbreak size. As a result, average costs per case calculated from these studies would substantially overestimate the cost per case if applied to larger outbreaks. We demonstrate the relationship between outbreak size and total cost in Figure 2, illustrating that small outbreaks have disproportionately high cost per case due to the initial large degree of fixed expenditures, while large outbreaks accumulate higher overall costs but at a substantially smaller cost per case.

**Table 3.** Marginal Cost per Case.

|  | Point Estimate | Robust Standard Error | 95% Confidence Interval |
| --- | --- | --- | --- |
| Fixed Cost ($\alpha$) | \$244,480.40 | \$113,943.06 | (\$21,152.01 - \$467,808.79) |
| Incremental Cost per Case ($\beta$) | \$16,197.13 | \$299.90 | (\$15,609.32 - \$16,784.94) |

**Figure 2.**
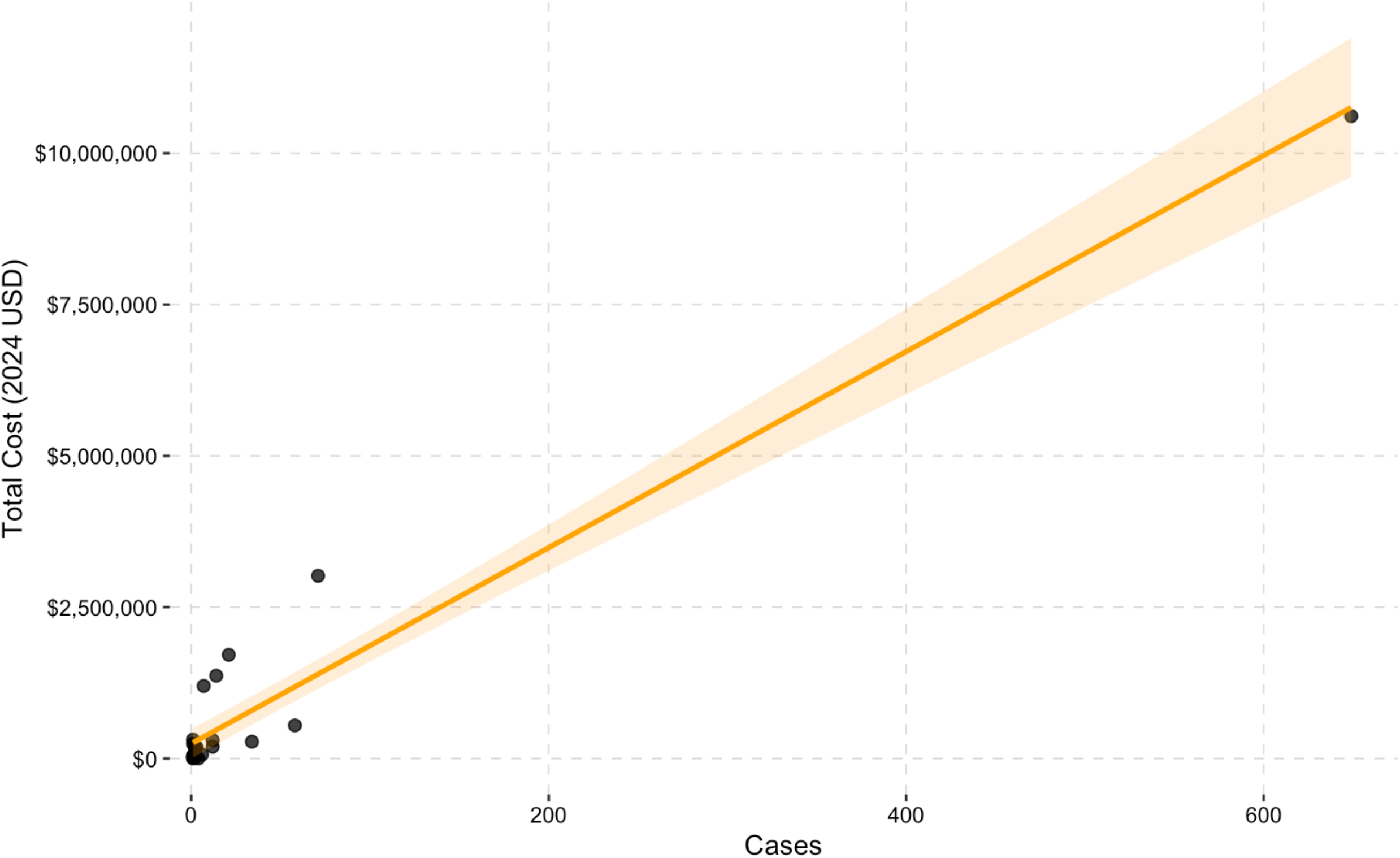
Total Outbreak Cost vs Number of Cases.

## Discussion

Our systematic review of measles outbreaks over the past 25 years in the United States demonstrates that these outbreaks impose a substantial financial and economic burden, regardless of outbreak size or setting. These findings build on Pike et al. 2020 ^3^ by incorporating more recent and larger outbreaks, including New York City (2018–2019), Washington State (2019), and the nationwide outbreak in 2025. Additionally, our review also included additional studies that were not presented in Pike et al. 2020, such as the Minneapolis outbreak by Hester et al. 2019 ^9^ and the US-bound Malaysian refugees’ study by Coleman et al. 2017.^8^ Coleman and colleagues reported event costs, including cost of labor, measles testing, vaccination, outpatient treatment, and hospitalization, in all states that involved refugees arriving at Los Angeles International Airport from Malaysia in 2011, even states without confirmed measles cases like Texas, New Hampshire, North Carolina, Maryland, Oklahoma, California, Wisconsin and Massachusetts, which is relevant to outbreak response cost estimates for the current nationwide outbreak. With the inclusion of these additional studies and the fact that more recent outbreaks were substantially larger than previous outbreaks, the overall costs in our analysis are higher than the costs reported by Pike et al. 2020. ^3^ In their review, the median total outbreak cost was approximately $152,308, ranging between $9,862 and $1,063,936. The median cost per case was estimated at $32,805 (range: $7,396– $76,154), while the median cost per contact was $223 (range: $81–$746).

The cost per case can be particularly high in outbreaks with fewer cases and contacts, especially in single-case events. This is due to large initial fixed costs associated with beginning an outbreak response, which are required immediately upon the first detection. These fixed costs include surveillance, measles testing systems, communication systems, and labor mobilization. However, as outbreak size expands, the incremental and marginal cost of additional cases due to an increase in contact tracing, measles vaccination campaigns, and direct medical costs and healthcare utilization incurred is typically lower than the cost per case of from a smaller outbreak. The new data sources from larger outbreaks enabled us to disaggregate incremental costs from fixed initiation costs as well as examine heterogeneity by costing perspective, adding to the usability of the economic evidence for both budgetary and impact forecasting for preparedness planning purposes by state and local governments as well as healthcare systems. Our regression analysis results demonstrate an average fixed cost of $244,480.40, while each additional measles case incurred an incremental cost of $16,197.13. For smaller outbreaks fixed outbreak response mobilization costs could contribute to greater than 50% of total costs incurred during an outbreak while for larger outbreaks they may contribute to less than 10% of total costs.

From a policy perspective, our results affirm that maintaining high measles vaccination coverage remains as one of the most cost-saving public health investments as it minimizes the chances of incurring high outbreak response fixed costs in the first place through localized herd immunity. Additionally, by quantifying both the fixed and incremental costs of measles outbreaks in the U.S., policymakers, healthcare administrators, and those involved in the outbreak preparedness and budgetary planning processes can have better estimates of the financial resource needs in the face of outbreaks of various scales as well as a better understanding of the potential savings and economic cost associated with prevention. Additionally, this research allows for sufficient cost data that it can be better disaggregated to highlight who bears the relative financial and economic burden of an outbreak and to what magnitude.

This analysis does come with several limitations. First, the cost reporting across studies was heterogeneous, with inconsistent definition of costing perspective. While we tried to account for this heterogeneity by presenting estimates unique to each perspective, there were insufficient data across outbreak size, particularly from the societal perspective, to draw strong conclusions of how indirect costs scaled with outbreak size. Many studies also lacked sufficient details on cost breakdowns, which hinders the ability for federal, state, and local planners to benefit from the detailed financial information for use in budgetary planning. Second, while our gray literature search expanded data on number cases, the availability and transparency on costs data were significantly more limited in these public reports than in the peer-reviewed published literature. Third, there remain significant gaps in the literature regarding the societal cost of measles outbreaks, as only one study in our analysis included estimates of the indirect productivity loss borne by patients themselves. Most studies excluded indirect costs like lost productivity, caregiver time, and long-term sequelae, leading to an underestimation of the total cost incurred from societal perspective as a result of an outbreak. Lastly, as the 2025 outbreaks are still ongoing, reported figures may be preliminary and subject to revision as official cost data become available.

Future research should prioritize standardizing frameworks for outbreak cost reporting, including harmonization around key disaggregation of costs by standardized time period and across standardized categories such as capital, training, labor, and medical care. Additionally, ensuring the inclusion of both direct and indirect costs from a societal perspective or at least making clear who is bearing the burden of included costs, is important for budgetary planning and outbreak preparedness planning for different actors involved in the health system. Finally, developing economic models for outbreak preparedness and response planning could help estimate potential national, state level, and local level resource needs for outbreak response paired with models for risk assessment to promote evidence-based planning. These models could also be used to then demonstrate potential savings attributable federal, state, and local budgets as well as to different entities within a healthcare system from sustained preventative measures, such as case-surveillance systems and maintaining high levels of measles vaccination coverage.

## Conclusion

Despite measles elimination in the United States, outbreaks continue to occur, with 2025 having the highest number of cases since elimination. These outbreaks impose substantial costs on both national and state governments, healthcare providers, and society. The economic burden of measles outbreaks is driven by high fixed costs and incremental costs that scale as the number of cases increases. Having better data on how these costs are incurred and by whom can substantially improve outbreak response budgetary planning and demonstrate the financial and economic impact that outbreaks have on healthcare systems. Given the high degree of fixed costs involved in outbreak response, minimizing the frequency of outbreaks in addition to the size of outbreaks remain a critical public health priority. This can be achieved through sustaining and strengthening measles vaccination coverage as well as ensuring financial resources exist for enabling rapid mobilization for containment.

## Data Availability

All data produced in the present study are available upon reasonable request to the authors

