## Appendix for "Quantifying the Cost of Measles Outbreak in the U.S. and How Costs Scale with Outbreak Size"

### Table A1. Inclusion/ Exclusion Criteria

| Category | Inclusion | Exclusion |
|---|---|---|
| **Population** | Studies conducted in the United States or primarily reporting U.S. data | Non-U.S. setting |
| **Timeframe** | 2000-2025 | Pre-2000 only |
| **Disease** | Measles | Other diseases |
| **Outcomes** | Any cost relevant to:<br>• treatment/care (direct medical, inpatient/outpatient, complications, societal cost)<br>• outbreak response (contact tracing, isolation/quarantine, PPE, staff time, logistics) | Without costs or outbreak response data, opinion/editorials without extractable cost data |
| **Publication Type** | Peer-reviewed articles; official government or CDC reports, credible economic analyses | Non-peer-reviewed sources (except credible government/agency reports), abstracts without full data |
| **Language** | English | Non-English without translation resources |

### Table A2. Literature Search Strategy

| **Categories** | **Query** | **Results** |
|---|---|---|
| Disease | ("measles"[MeSH Terms] OR measles[tiab] OR rubella[tiab]) | 39,903 |
| Cost/ Economic Outcomes | ( "cost of illness"[MeSH Terms] OR "costs and cost analysis"[MeSH Terms] OR "health care costs"[MeSH Terms] OR cost*[tiab] OR economic*[tiab] OR financial*[tiab] OR expenditure*[tiab] OR budget[tiab] ) | 1,531,991 |

| | | |
|---|---|---|
| Intervention/ Outbreak Response | ( "vaccination"[MeSH Terms] OR "immunization"[MeSH Terms] OR vaccine[tiab] OR "public health campaign"[tiab] OR "public health messaging"[tiab] OR "outbreak response"[tiab] OR "contact tracing"[MeSH Terms] OR "personal protective equipment"[MeSH Terms] OR containment[tiab] OR quarantine[MeSH Terms] OR isolation[MeSH Terms] OR hospitalization[MeSH Terms] OR treatment[tiab] OR management[tiab] OR review[tiab] OR "systematic review"[tiab] ) | 9,272,547 |
| Countries | (("United States"[MeSH Terms] OR "United States"[tiab] OR "U.S."[tiab] OR USA[tiab] OR Alabama[MeSH Terms] OR Alabama[TIAB] OR Alaska[MeSH Terms] OR Alaska[TIAB] OR Arizona[MeSH Terms] OR Arizona[TIAB] OR Arkansas[MeSH Terms] OR Arkansas[TIAB] OR California[MeSH Terms] OR California[TIAB] OR Colorado[MeSH Terms] OR Colorado[TIAB] OR Connecticut[MeSH Terms] OR Connecticut[TIAB] OR Delaware[MeSH Terms] OR Delaware[TIAB] OR Florida[MeSH Terms] OR Florida[TIAB] OR Georgia[MeSH Terms] OR Georgia[TIAB] OR Hawaii[MeSH Terms] OR Hawaii[TIAB] OR Idaho[MeSH Terms] OR Idaho[TIAB] OR Illinois[MeSH Terms] OR Illinois[TIAB] OR Indiana[MeSH Terms] OR Indiana[TIAB] OR Iowa[MeSH Terms] OR Iowa[TIAB] OR Kansas[MeSH Terms] OR Kansas[TIAB] OR Kentucky[MeSH Terms] OR Kentucky[TIAB] OR Louisiana[MeSH Terms] OR Louisiana[TIAB] OR Maine[MeSH Terms] OR Maine[TIAB] OR Maryland[MeSH Terms] OR Maryland[TIAB] OR Massachusetts[MeSH Terms] OR Massachusetts[TIAB] OR Michigan[MeSH Terms] OR Michigan[TIAB] OR Minnesota[MeSH Terms] OR Minnesota[TIAB] OR Mississippi[MeSH Terms] OR Mississippi[TIAB] OR Missouri[MeSH Terms] OR Missouri[TIAB] OR Montana[MeSH Terms] OR Montana[TIAB] OR Nebraska[MeSH Terms] OR Nebraska[TIAB] OR Nevada[MeSH Terms] OR Nevada[TIAB] OR "New Hampshire"[MeSH Terms] OR "New Hampshire"[TIAB] OR "New Jersey"[MeSH Terms] OR "New Jersey"[TIAB] OR "New Mexico"[MeSH Terms] OR "New Mexico"[TIAB] OR "New York"[MeSH Terms] OR "New York"[TIAB] OR "North Carolina"[MeSH Terms] OR "North Carolina"[TIAB] OR "North Dakota"[MeSH Terms] OR "North Dakota"[TIAB] OR Ohio[MeSH Terms] OR Ohio[TIAB] OR Oklahoma[MeSH Terms] OR Oklahoma[TIAB] OR Oregon[MeSH Terms] OR Oregon[TIAB] OR Pennsylvania[MeSH Terms] OR Pennsylvania[TIAB] OR "Rhode Island"[MeSH Terms] OR "Rhode Island"[TIAB] OR "South Carolina"[MeSH Terms] OR "South Carolina"[TIAB] OR "South Dakota"[MeSH Terms] OR "South Dakota"[TIAB] OR Tennessee[MeSH Terms] OR Tennessee[TIAB] OR Texas[MeSH Terms] OR Texas[TIAB] OR Utah[MeSH Terms] OR Utah[TIAB] OR Vermont[MeSH Terms] OR Vermont[TIAB] OR Virginia[MeSH | 2,026,053 |

| | | |
|---|---|---|
| | Terms] OR Virginia[TIAB] OR Washington[MeSH Terms] OR Washington[TIAB] OR "West Virginia"[MeSH Terms] OR "West Virginia"[TIAB] OR Wisconsin[MeSH Terms] OR Wisconsin[TIAB] OR Wyoming[MeSH Terms] OR Wyoming[TIAB])) | |
| Timeline | (2000:2025[pdat]) | 120 |

**Table A3. Measles outbreak data extraction results from Gray Literature.**

| *Ref* | *Citation* | *Year* | *Location* | *Number of Cases* | *Total cost (USD)* |
|---|---|---|---|---|---|
| *1* | Measles outbreak - California, December 2014–February 2015. Centers for Disease Control and Prevention. February 20, 2015. https://www.cdc.gov/mmwr/preview/mmwrhtml/mm6406a5.htm. | 2014-2015 | California | 110 | |
| *2* | Measles outbreak - California, December 2014–February 2015. Centers for Disease Control and Prevention. February 20, 2015. https://www.cdc.gov/mmwr/preview/mmwrhtml/mm6406a5.htm. | 2014-2015 | Arizona | 7 | |
| *3* | Measles outbreak - California, December 2014–February 2015. Centers for Disease Control and Prevention. February 20, 2015. https://www.cdc.gov/mmwr/preview/mmwrhtml/mm6406a5.htm. | 2014-2015 | Colorado | 1 | |
| *4* | Measles outbreak - California, December 2014–February 2015. Centers for Disease Control and Prevention. February 20, 2015. https://www.cdc.gov/mmwr/preview/mmwrhtml/mm6406a5.htm. | 2014-2015 | Nebraska | 1 | |
| *5* | Measles outbreak - California, December 2014–February 2015. Centers for Disease Control and Prevention. February 20, 2015. https://www.cdc.gov/mmwr/preview/mmwrhtml/mm6406a5.htm. | 2014-2015 | Oregon | 1 | |
| *6* | Measles outbreak - California, December 2014–February 2015. Centers for Disease Control and Prevention. February 20, 2015. https://www.cdc.gov/mmwr/preview/mmwrhtml/mm6406a5.htm. | 2014-2015 | Utah | 3 | |
| *7* | Measles outbreak - California, December 2014–February 2015. Centers for Disease Control and Prevention. February 20, 2015. https://www.cdc.gov/mmwr/preview/mmwrhtml/mm6406a5.htm. | 2014-2015 | Washington | 2 | |
| *8* | Oakland County measles outbreak response. Oakland County Measles Outbreak Response Oakland County Health Department Success Story. 2019. https://www.michigan.gov/mdhhs/-/media/Project/Websites/mdhhs/Keeping-Michigan-Healthy/Local-Health-Services/02_PHHS-Block-Grant/Success-Stories/MI-LHD-Success-Story-Oakland-County-Measles.pdf?rev=9174bbf702904c9386a5eb40a0dbb599&hash=2DA5413CEC7C2FDE2D8BD31B3FD9D022. | 2019 | Michigan | 44 | |
| *9* | Notes from the field: Measles outbreaks from imported cases in Orthodox Jewish communities - New York and New Jersey, 2018–2019. Centers for Disease Control and Prevention. May 16, 2019. https://www.cdc.gov/mmwr/volumes/68/wr/mm6819a4.htm. | 2018-2019 | Rockland County, NY | 242 | |
| *10* | Measles cases and outbreaks. Centers for Disease Control and Prevention. September 3, 2025. https://www.cdc.gov/measles/data-research/index.html. | 2025 | Alabama | 1 | |

| | | | | |
|---|---|---|---|---|
| *11* | Measles cases and outbreaks. Centers for Disease Control and Prevention. September 3, 2025. https://www.cdc.gov/measles/data-research/index.html. | 2025 | Alaska | 3 |
| *12* | Measles cases and outbreaks. Centers for Disease Control and Prevention. September 3, 2025. https://www.cdc.gov/measles/data-research/index.html. | 2025 | Arizona | 28 |
| *13* | Measles cases and outbreaks. Centers for Disease Control and Prevention. September 3, 2025. https://www.cdc.gov/measles/data-research/index.html. | 2025 | Arkansas | 8 |
| *14* | 1. *Measles cases and outbreaks. Centers for Disease Control and Prevention. September 3, 2025. https://www.cdc.gov/measles/data-research/index.html.*<br>2. *Health D of P. Measles. September 2, 2025. https://www.cdph.ca.gov/Programs/CID/DCDC/Pages/Immunization/measles.aspx.* | 2025 | California | 20 |
| *15* | Measles cases and outbreaks. Centers for Disease Control and Prevention. September 3, 2025. https://www.cdc.gov/measles/data-research/index.html. | 2025 | Colorado | 21 |
| *16* | Measles cases and outbreaks. Centers for Disease Control and Prevention. September 3, 2025. https://www.cdc.gov/measles/data-research/index.html. | 2025 | Florida | 6 |
| *17* | Measles cases and outbreaks. Centers for Disease Control and Prevention. September 3, 2025. https://www.cdc.gov/measles/data-research/index.html. | 2025 | Georgia | 6 |
| *18* | Measles cases and outbreaks. Centers for Disease Control and Prevention. September 3, 2025. https://www.cdc.gov/measles/data-research/index.html. | 2025 | Hawaii | 2 |
| *19* | Measles cases and outbreaks. Centers for Disease Control and Prevention. September 3, 2025. https://www.cdc.gov/measles/data-research/index.html. | 2025 | Idaho | 3 |
| *20* | Measles cases and outbreaks. Centers for Disease Control and Prevention. September 3, 2025. https://www.cdc.gov/measles/data-research/index.html. | 2025 | Illinois | 10 |
| *21* | Measles cases and outbreaks. Centers for Disease Control and Prevention. September 3, 2025. https://www.cdc.gov/measles/data-research/index.html. | 2025 | Indiana | 8 |
| *22* | Measles cases and outbreaks. Centers for Disease Control and Prevention. September 3, 2025. https://www.cdc.gov/measles/data-research/index.html. | 2025 | Iowa | 8 |
| *23* | Measles cases and outbreaks. Centers for Disease Control and Prevention. September 3, 2025. https://www.cdc.gov/measles/data-research/index.html. | 2025 | Kansas | 90 |

| | | | | |
|---|---|---|---|---|
| 24 | Measles cases and outbreaks. Centers for Disease Control and Prevention. September 3, 2025. https://www.cdc.gov/measles/data-research/index.html. | 2025 | Kentucky | 13 |
| 25 | Measles cases and outbreaks. Centers for Disease Control and Prevention. September 3, 2025. https://www.cdc.gov/measles/data-research/index.html. | 2025 | Louisiana | 2 |
| 26 | Measles cases and outbreaks. Centers for Disease Control and Prevention. September 3, 2025. https://www.cdc.gov/measles/data-research/index.html. | 2025 | Maryland | 3 |
| 27 | Measles cases and outbreaks. Centers for Disease Control and Prevention. September 3, 2025. https://www.cdc.gov/measles/data-research/index.html. | 2025 | Michigan | 27 |
| 28 | Measles cases and outbreaks. Centers for Disease Control and Prevention. September 3, 2025. https://www.cdc.gov/measles/data-research/index.html. | 2025 | Minnesota | 5 |
| 29 | Measles cases and outbreaks. Centers for Disease Control and Prevention. September 3, 2025. https://www.cdc.gov/measles/data-research/index.html. | 2025 | Missouri | 6 |
| 30 | Measles cases and outbreaks. Centers for Disease Control and Prevention. September 3, 2025. https://www.cdc.gov/measles/data-research/index.html. | 2025 | Montana | 31 |
| 31 | Measles cases and outbreaks. Centers for Disease Control and Prevention. September 3, 2025. https://www.cdc.gov/measles/data-research/index.html. | 2025 | Nebraska | 1 |
| 32 | Measles cases and outbreaks. Centers for Disease Control and Prevention. September 3, 2025. https://www.cdc.gov/measles/data-research/index.html. | 2025 | New Jersey | 10 |
| 33 | 1. *Measles cases and outbreaks. Centers for Disease Control and Prevention. September 3, 2025. https://www.cdc.gov/measles/data-research/index.html.*<br>2. *New Mexico reports 100 measles cases. August 14, 2025. https://www.nmhealth.org/news/awareness/2025/8/?view=2262.* | 2025 | New Mexico | 100 |
| 34 | Measles cases and outbreaks. Centers for Disease Control and Prevention. September 3, 2025. https://www.cdc.gov/measles/data-research/index.html. | 2025 | New York | 7 |
| 35 | Measles cases and outbreaks. Centers for Disease Control and Prevention. September 3, 2025. https://www.cdc.gov/measles/data-research/index.html. | 2025 | New York City | 8 |
| 36 | Measles cases and outbreaks. Centers for Disease Control and Prevention. September 3, 2025. https://www.cdc.gov/measles/data-research/index.html. | 2025 | North Dakota | 36 |
| 37 | Measles cases and outbreaks. Centers for Disease Control and Prevention. September 3, 2025. https://www.cdc.gov/measles/data-research/index.html. | 2025 | Ohio | 30 |

| | | | | | |
|---|---|---|---|---|---|
| *38* | Measles cases and outbreaks. Centers for Disease Control and Prevention. September 3, 2025. https://www.cdc.gov/measles/data-research/index.html. | 2025 | Oklahoma | 17 | |
| *39* | Measles cases and outbreaks. Centers for Disease Control and Prevention. September 3, 2025. https://www.cdc.gov/measles/data-research/index.html. | 2025 | Oregon | 1 | |
| *40* | Measles cases and outbreaks. Centers for Disease Control and Prevention. September 3, 2025. https://www.cdc.gov/measles/data-research/index.html. | 2025 | Pennsylvani a | 16 | |
| *41* | Measles cases and outbreaks. Centers for Disease Control and Prevention. September 3, 2025. https://www.cdc.gov/measles/data-research/index.html. | 2025 | Rhode Island | 1 | |
| *42* | Measles cases and outbreaks. Centers for Disease Control and Prevention. September 3, 2025. https://www.cdc.gov/measles/data-research/index.html. | 2025 | South Carolina | 2 | |
| *43* | Measles cases and outbreaks. Centers for Disease Control and Prevention. September 3, 2025. https://www.cdc.gov/measles/data-research/index.html. | 2025 | South Dakota | 12 | |
| *44* | Measles cases and outbreaks. Centers for Disease Control and Prevention. September 3, 2025. https://www.cdc.gov/measles/data-research/index.html. | 2025 | Tennessee | 6 | |
| *45* | 1. *Measles outbreak – August 12, 2025. Measles Outbreak – August 12, 2025 \| Texas DSHS. August 12, 2025. https://www.dshs.texas.gov/news-alerts/measles-outbreak-2025.*<br>2. *Langford T. State health officials declare West Texas measles outbreak over. The Texas Tribune. August 18, 2025. https://www.texastribune.org/2025/08/18/texas-measles-west-outbreak-over-dshs/.*<br>3. *Measles cases and outbreaks. Centers for Disease Control and Prevention. September 3, 2025. https://www.cdc.gov/measles/data-research/index.html.* | 2025 | Texas | 802 | $10 million |
| *46* | Measles cases and outbreaks. Centers for Disease Control and Prevention. September 3, 2025. https://www.cdc.gov/measles/data-research/index.html. | 2025 | Utah | 15 | |
| *47* | Measles cases and outbreaks. Centers for Disease Control and Prevention. September 3, 2025. https://www.cdc.gov/measles/data-research/index.html. | 2025 | Vermont | 2 | |
| *48* | Measles cases and outbreaks. Centers for Disease Control and Prevention. September 3, 2025. https://www.cdc.gov/measles/data-research/index.html. | 2025 | Virginia | 3 | |
| *49* | Measles cases and outbreaks. Centers for Disease Control and Prevention. September 3, 2025. https://www.cdc.gov/measles/data-research/index.html. | 2025 | Washington | 10 | |

| | | | | |
|---|---|---|---|---|
| *50* | Measles cases and outbreaks. Centers for Disease Control and Prevention. September 3, 2025. https://www.cdc.gov/measles/data-research/index.html. | 2025 | Wisconsin | 24 |
| *51* | Measles cases and outbreaks. Centers for Disease Control and Prevention. September 3, 2025. https://www.cdc.gov/measles/data-research/index.html. | 2025 | Wyoming | 9 |
